# Clozapine treatment and risk of COVID-19

**DOI:** 10.1101/2020.06.17.20133595

**Authors:** Risha Govind, Daniela Fonseca de Freitas, Megan Pritchard, Richard D. Hayes, James H. MacCabe

## Abstract

**Background:** Clozapine, an antipsychotic with unique efficacy in treatment resistant psychosis, is associated with increased susceptibility to infection, including pneumonia.

**Aims:** To investigate associations between clozapine treatment and increased risk of COVID-19 in patients with schizophrenia-spectrum disorders who are receiving antipsychotic medications, using electronic health records data, in a geographically defined population in London.

**Method:** Using information from South London and Maudsley NHS Foundation Trust (SLAM) clinical records, via the Clinical Record Interactive Search system, we identified 6,309 individuals who had an ICD-10 diagnosis of schizophrenia-spectrum disorders and were taking antipsychotics at the time on the COVID-19 pandemic onset in the UK. People who were on clozapine treatment were compared with those on any other antipsychotic treatment for risk of contracting COVID-19 between 1 March and 18 May 2020. We tested associations between clozapine treatment and COVID-19 infection, adjusting for gender, age, ethnicity, BMI, smoking status, and SLAM service use.

**Results:** Of 6,309 patients, 102 tested positive for COVID-19. Individuals who were on clozapine had increased risk of COVID-19 compared with those who were on other antipsychotic medication (unadjusted HR = 2.62 (95% CI 1.73 - 3.96), which was attenuated after adjusting for potential confounders, including clinical contact (adjusted hazard ratio HR=1.76, 95% CI 1.14 - 2.72).

**Conclusions:** These findings provide support for the hypothesis that clozapine treatment is associated with an increased risk of COVID-19. Further research will be needed in other samples to confirm this association. Potential clinical implications are discussed.

## INTRODUCTION

Clozapine is an antipsychotic with unique efficacy in treatment resistant psychosis, and, for many patients, it is the only effective treatment (1). It is associated with a reduction in hospital admissions, overall mortality, and suicide risk in schizophrenia (2–5). Patients with schizophrenia have an increased mortality compared with the general population (6,7). Some of this excess mortality is attributable to pneumonia (8–11). Much of this increase may be attributable to higher rates of smoking in schizophrenia patients (12). However, there appears to be an additional effect of clozapine treatment (13–16). In the study of Kuo and colleagues, treatment with clozapine was associated with approximately a doubling of the risk of pneumonia among patients with schizophrenia (14). However, confounding by indication could have affected these results: clozapine is prescribed to treatment resistant patients, and such patients are likely to suffer a range of comorbidities that increase the risk of infection, such as smoking and other substance misuse, poor diet and a sedentary lifestyle (17). It is also plausible that some of the adverse effects of clozapine, such as diabetes, weight gain, or hypersalivation (leading to aspiration pneumonia (18)) could lie on the causal pathway between clozapine treatment and the risk of infection. Clozapine treatment appears to have multiple effects on the innate immune system, including transient eosinophilia, cytokine release and fever during early treatment, and neutropenia and agranulocytosis in a small minority (19). There is emerging evidence that adaptive immunity is also affected by clozapine, (20) with a reduction in all three classes of circulating immunoglobulins (IgM, IgA, and IgG) in clozapine-treated patients compared with those on other antipsychotics. COVID-19 is a novel infection caused by SARS-Cov2, causing pneumonia in severe cases, which arose in China in late 2019 and was declared a global pandemic by the WHO in March 2020 (21). Given the effects of clozapine on immunity and the increased risk of pneumonia, we investigated whether clozapine treatment was associated with an increased risk of COVID-19 in patients with schizophrenia and other psychoses treated with antipsychotics in a geographically defined population in London during the COVID-19 pandemic.

## METHOD

### Setting and Ethics Statement

This retrospective cohort study used data from the South London and Maudsley NHS Foundation Trust (SLAM), one of Europe’s largest secondary mental healthcare providers. In the United Kingdom, mental health services are provided based on defined geographical catchment areas under the National Health Service (NHS). SLAM provides all aspects of secondary mental health care to over 1.3 million people of four London boroughs (Lambeth, Southwark, Lewisham, and Croydon). From 2006, SLAM has used a fully electronic health records system, and the Clinical Records Interactive Search system (CRIS), supported by NIHR Specialist Biomedical Research Centre for Mental Health. CRIS was established in 2008 to enable researchers to search and retrieve de-identified clinical records from SLAM. The protocol for CRIS has been described in detail in an open-access publication (22). CRIS was approved as an anonymised data resource for secondary analysis by Oxfordshire Research Ethics Committee C (reference 18/SC/0372). The data linkage to King’s College Hospital for admissions regarding COVID-19 infections took place under Regulation 3(2) and Regulation 3(3) of the Health Service Control of Patient Information Regulations 2002 (COPI).

### Analytical Cohort & Data Extraction

The cohort comprised of individuals who filled all 3 of the following inclusion criteria: ICD-10, diagnosis of schizophrenia-spectrum disorders (F2*); taking antipsychotic medication between December 1, 2019 and March 1, 2020; and receiving outpatient or inpatient care at SLAM on March 1, 2020. This date was chosen because it was before March 11, 2020, the date of the first diagnosed case on COVID-19 in SLAM, so there was no risk of reverse causation (the presence of COVID-19 infection affecting the exposures).

SQL Server Management Studio version 15.0 was used to extract the data. The day of data extraction was May 18, 2020. The index period, from which medication data were gathered, was from December 1, 2019 to March 1, 2020. Patients were followed-up from March 1, 2020 until they were diagnosed COVID-19 positive, died, or reached the end of the observation period, May 18, 2020, which ever occurred first.

The data extracted from CRIS was of two types: structured fields, entered by the clinicians and administrators, and data extracted from free-text fields of clinical records, using custom-built Natural Language Processing (NLP) algorithms (23). NLP algorithms are able to outperform key word searches because they take into account the linguistic context around terms of interest, for example, temporal modifiers (e.g., “on clozapine” versus “previously took clozapine”). Data from four NLP algorithms were used in this study, diagnosis, medication, smoking and body mass index (BMI).

The diagnosis algorithm was used for the inclusion criteria to identify individuals who were ever diagnosed with ICD-10 diagnoses of schizophrenia-spectrum disorders (F2*). The overall precision and recall scores for the diagnosis algorithm are 100% and 65% respectively (23).

The medication algorithm data was also used for the inclusion criteria to identify individuals who were on an antipsychotic medication between December 1, 2019, and March 1, 2020, the index period. This algorithm provides specific results for antipsychotic medication. The antipsychotic prescriptions included in this analysis study were Clozapine, Olanzapine, Risperidone, Aripiprazole, Amisulpride, Paliperidone, Flupentixol, Haloperidol, Zuclopenthixol, Quetiapine, Fluphenazine, Piportil, Sulpiride, Lurasidone, Trifluoperazine, Chlorpromazine, Pipotiazine, Penfluridol, Droperidol, Pimozide, Thioridazine, Promazine, Ziprasidone Hydrochloride, Levomepromazine and Pericyazine. The precision and recall scores for the antipsychotics part of the medication algorithm are 88% and 90%, respectively (23).

The smoking algorithm was used to identify the smoking status of each patient. The “current smoker” status was based on data from dates between March 1, 2019 and March 1, 2020. The “past smoker” and “never smoked” status were based on all available information in the electronic health record. In the underlying patient records, smoking status may be recorded repeatedly, i.e., each time this information is entered into the patient record. Consequently, for some patients, the smoking algorithm may identify more than one smoking status per patient. Where this was the case, we took the highest smoking status in the hierarchy “current smoker” > “past smoker” >“never smoked”. The precision (P) and recall (R) scores for each status of the Smoking algorithm are as follows: for “current smoker” status, P=79% and R=87%; for “past smoker” status, P=68% and R=38%; for “never smoked” status, P=72% and R=75% (23).

The BMI algorithm was used to extract the most recent BMI of each patient in the entire patient record. To exclude erroneous values from the results of this algorithm, we rejected values outside the range 15 - 70. The overall precision and recall scores for the BMI algorithm are 89% and 78%, respectively (23).

All the NLP algorithms outputs were also supplemented by the data in the structured fields, data in the health records, such as, data from ICD 10 diagnosis forms for diagnosis data, and pharmacy dispensary data for medication data.

Of all patients in SLAM, 6309 patients met the inclusion criteria of individuals with ICD-10 diagnoses of schizophrenia-spectrum disorders (F2*) who were on antipsychotic medication during the index period.

### Main outcome measure

The outcome of interest was infection with COVID-19 during the follow-up period (March 1, 2020 to May 18, 2020). These data were collated by combining information from the SLAM pathology lab results data, the presence of a clinician-entered alert on SLAM records: “COVID-19 positive” and information provided by local general hospitals (King’s College Hospital and Princess Royal University Hospital) for COVID-19 related admissions.

### Exposure variables

In keeping with the cohort study design, all exposures were recorded prior to the start of follow up. People who were on clozapine treatment at any time between December 1, 2019 and March 1, 2020, were designated as the exposed group. Those on any type or combination of antipsychotic treatment that did not include clozapine during this time constituted the unexposed group.

We included in our analysis individual-level information on sociodemographic characteristics, health, and service use. The sociodemographic information was age, gender, and ethnicity. The health information was smoking status and BMI. The SLAM services use information were data on whether the patient was an inpatient on March 1, 2020, and the number of days the patient was in contact with the SLAM services between December 1, 2019 and March 1, 2020. The contact with SLAM services included any form of inpatient and outpatient communication such as email or phone or face-to-face consultations.

### Statistical analysis

The data were analysed using STATA for Windows version 15.1. Using Cox proportional hazard models, we calculated hazard ratios for COVID-19 positive status, in clozapine treated patients versus those treated with other antipsychotics. We censored observations at the date of death, date of COVID-19 positive test, or May 18, 2020, whichever occurred first. We confirmed that the data satisfied the proportional-hazards assumptions using Schoenfeld residuals.

Three variables contained missing data: smoking, ethnicity, and BMI. Firstly, we analysed the entire cohort using only variables with no missing data. Then, in a complete case analysis, we ran the same analyses, excluding individuals for whom there were any missing data across any of the exposures investigated (n=5,535). Since the results were very similar, we were confident that the complete case analysis was unlikely to suffer from undue selection bias. We have presented results from the complete case analysis and have included the results of the whole cohort analysis in the supplementary material.

Crude and adjusted models were constructed, first controlling for age, gender, and ethnicity; then controlling for age, gender, ethnicity, inpatient status and number of contact days with the SLAM services; lastly, a fully adjusted model controlling for all variables, age, gender, ethnicity, inpatient status, number of contact days with the SLAM services smoking status and BMI. All models were built using data from the 5,535 individuals with complete data. The above models were repeated using the whole cohort (n=6,309) without including the variables with missing data (ethnicity, smoking status, BMI), these results are in supplementary table 1.

**Table 1:** Sample Characteristics of all SLAM patients who qualified for the inclusion criteria, presented according to those who were and were not on clozapine-treatment

|  | Not on clozapine-treatment<br>n (%) | On clozapine treatment<br>n (%) |
| --- | --- | --- |
| Total (n=6,309) | 5,027 (79.68) | 1,282 (20.32) |
| Gender |  |  |
| Male | 3,047 (60.61) | 847 (66.07) |
| Female | 1,980 (39.39) | 435 (33.93) |
| Age |  |  |
| < 29 years | 817 (16.25) | 131 (10.22) |
| 30 to < 40 years | 1,055 (20.99) | 281 (21.92) |
| 40 to < 50 years | 1,082 (21.52) | 338 (26.37) |
| 50 to < 60 years | 1,145 (22.78) | 369 (28.78) |
| 60 to < 70 years | 546 (10.86) | 127 (9.91) |
| 70 years and over | 382 (7.60) | 36 (2.81) |
| Ethnicity |  |  |
| White | 1,579 (31.41) | 516 (40.25) |
| Black | 2,607 (51.86) | 586 (45.71) |
| Asian, Other & Not Stated | 691 (13.75) | 170 (13.26) |
| Missing | 150 (2.98) | 10 (0.78) |
| Inpatient on the first day of follow-up period* |  |  |
| No | 4,735 (94.19) | 1,113 (86.82) |
| Yes | 292 (5.81) | 169 (13.18) |
| SLAM service contact during index period** |  |  |
| < 4 days | 1,902 (37.84) | 242 (18.88) |
| 4 to 7 days | 1,575 (31.33) | 479 (37.36) |
| 8 days and more | 1,550 (30.83) | 561 (43.76) |
| Smoking status |  |  |
| Current smoker | 3,154 (62.74) | 1,028 (80.19) |
| Past smoker | 1,457 (28.98) | 232 (18.10) |
| Never smoked | 326 (6.48) | 18 (1.40) |
| Missing | 90 (1.79) | 4 (0.31) |
| BMI |  |  |
| Underweight & healthy | 1,601 (31.85) | 264 (20.59) |
| Overweight | 1,335 (26.56) | 358 (27.93) |
| Obese | 1,499 (29.82) | 610 (47.58) |
| Missing | 592 (11.78) | 50 (3.90) |

\*: first date of follow period is March 1, 2020
\*\*: index period is between December 1, 2019 and March 1, 2020, which is 3 months prior to the follow-up period

## RESULTS

There were 6,309 active patients with schizophrenia-spectrum disorders (F2*) who were receiving any type of antipsychotic treatment during the beginning of the follow-up period. The sample mean age was 46.5 (SD=14.8), and men account for 61.7% of the sample. Sample’s ethnic description, 33.2% White (White British; Irish or any other White Background), 50.6% Black (including Black African, Black Caribbean, Other Black background, White and African, and White and Caribbean), 13.7% any Asian and Other ethnic background, 2.5% had missing data on ethnicity.

Table 1 summarises the demographic features of all the SLAM patients who qualified for the inclusion criteria (n=6309). Of the individuals who were on clozapine, 66% were male, 46% were Black, 80% were current smokers, and 48% had high BMI (obese). Compared to patients not on clozapine treatment, a higher proportion of clozapine treated patients were inpatients in the hospital on March 1, 2020 (13% vs 6%), and clozapine patients had more contact days with the SLAM services in the prior 3 months.

Table 2 summarises the demographic features presented according to their outcome status, COVID-19 positive or not COVID-19 positive. Of those who were COVID-19 positive, 41% were receiving clozapine treatment, whereas those who were not COVID-19 positive, only 20% were receiving clozapine treatment. A higher proportion of COVID-19 positive patients were inpatients and COVID-19 positive patients had more contact days with the SLAM services

**Table 2:** Sample characteristics of all SLAM patients who qualified for the inclusion criteria, presented according to those who tested positive for COVID-19 and those who did not, during the follow-up period (March 1, 2020 until May 18, 2020, inclusive).

|  | Not COVID-19<br>Positive<br>n (%) | COVID-19<br>Positive<br>n (%) |
| --- | --- | --- |
| Total sample (n=6,309) | 6,207 (98.38) | 102 (1.62) |
| On clozapine treatment |  |  |
| No | 4,967 (80.02) | 60 (58.82) |
| Yes | 1,240 (19.98) | 42 (41.18) |
| Gender |  |  |
| Male | 3,838 (61.83) | 56 (54.90) |
| Female | 2,369 (38.17) | 46 (45.10) |
| Age |  |  |
| < 29 years | 937 (15.10) | 11 (10.78) |
| 30 to 39 years | 1,315 (21.19) | 21 (20.59) |
| 40 to 49 years | 1,408 (22.68) | 12 (11.76) |
| 50 to 59 years | 1,485 (23.92) | 29 (28.43) |
| 60 to 69 years | 657 (10.58) | 16 (15.69) |
| 70 years and over | 405 (6.52) | 13 (12.75) |
| Ethnicity |  |  |
| White | 2,069 (33.33) | 26 (25.49) |
| Black | 3,129 (50.41) | 64 (62.75) |
| Asian, Other & Not Stated | 853 (13.74) | 8 (7.84) |
| Missing | 156 (2.51) | 4 (3.92) |
| Inpatient on the first day of follow-up period* |  |  |
| No | 5,804 (93.51) | 44 (43.14) |
| Yes | 403 (6.49) | 58 (56.86) |
| SLAM service contact during index period** |  |  |
| < 4 days | 2,137 (34.43) | 7 (6.86) |
| 4 to 7 days | 2,035 (32.79) | 19 (18.63) |
| 8 days and more | 2,035 (32.79)) | 76 (74.51) |
| Smoking status |  |  |
| Current smoker | 4,104 (66.12) | 78 (76.47) |
| Past smoker | 1,669 (26.89) | 20 (19.61) |
| Never smoked | 342 (5.51) | 2 (1.96) |
| Missing | 92 (1.48) | 2 (1.96) |
| BMI |  |  |
| Underweight & healthy | 1,843 (29.69) | 22 (21.57) |
| Overweight | 1,675 (26.99) | 18 (17.65) |
| Obese | 2,054 (33.09) | 55 (53.92) |
| Missing | 635 (10.23) | 7 (6.86) |

\*: first date of follow period is March 1, 2020
\*\* : index period is between December 1, 2019 and March 1, 2020, which is 3 months prior to the follow-up period

The Cox regression analysis was performed with data of 5,535 individuals with complete information (774 participants excluded due to missing data, see details in Table 1), and the mean (SD) follow-up period was 78.00 (7.03) days. Of these 5,535 individuals, 92 tested positive for infection with COVID-19 during the follow-up period. Table 3 shows the hazard ratio for COVID-19 associated with being on clozapine-treatment in the crude and adjusted model. The crude model shows a hazard ratio (HR) of 2.62 (95% CI 1.73 - 3.96) for patients receiving clozapine-treatment and COVID-19 positive. This increased to 3.06 (95% CI 2.01 – 4.67) after adjusting for sociodemographic factors (age, gender, ethnicity). It was attenuated to 1.85 (95% CI 1.20 – 2.85) after adjusting for inpatient status and SLAM service contact. It was further attenuated to 1.76 (95% CI 1.14 – 2.72) after adjusting for BMI and smoking status.

**Table 3:** Multivariate Cox analysis of association between receiving clozapine treatment and COVID-19 infection between March 1, 2020 until May 18, 2020, inclusive in 5 535 individuals (92 COVID-19 Positive Cases).

| Risk Factors | Hazard Ratio<br>(95% CI) | Hazard Ratio<br>(95% CI) | Hazard Ratio<br>(95% CI) | Hazard Ratio<br>(95% CI) |
| --- | --- | --- | --- | --- |
| On clozapine treatment |  |  |  |  |
| No | 1.00 | 1.00 | 1.00 | 1.00 |
| Yes | 2.62 (1.73 - 3.96) | 3.06 (2.01 - 4.67) | 1.85 (1.20 - 2.85) | 1.76 (1.14 - 2.72) |
| Gender |  |  |  |  |
| Male |  | 1.00 | 1.00 | 1.00 |
| Female |  | 1.30 (0.86 - 1.97) | 1.43 (0.94 - 2.17) | 1.32 (0.86 - 2.04) |
| Age |  |  |  |  |
| < 29 years |  | 1.00 | 1.00 | 1.00 |
| 30 to 39 years |  | 1.03 (0.49 - 2.18) | 1.25 (0.59 - 2.66) | 1.14 (0.53 - 2.42) |
| 40 to 49 years |  | 0.54 (0.23 - 1.27) | 0.69 (0.29 - 1.63) | 0.64 (0.27 - 1.52) |
| 50 to 59 years |  | 1.30 (0.64 - 2.64) | 1.95 (0.95 - 3.98) | 1.78 (0.87 - 3.64) |
| 60 to 69 years |  | 1.73 (0.78 - 3.83) | 2.88 (1.29 - 6.45) | 2.76 (1.22 - 6.23) |
| 70 years and over |  | 3.31 (1.43 - 7.66) | 4.10 (1.76 - 9.51) | 4.35 (1.85 - 10.26) |
| Ethnicity |  |  |  |  |
| White |  | 1.00 | 1.00 | 1.00 |
| Black |  | 1.98 (1.23 - 3.19) | 1.81 (1.12 - 2.91) | 1.70 (1.04 - 2.77) |
| Asian, Other, Not Stated |  | 0.74 (0.30 - 1.82) | 0.75 (0.31 - 1.84) | 0.78 (0.32 - 1.92) |
| Inpatient on the first day<br>of follow-up period* |  |  |  |  |
| No |  |  | 1.00 | 1.00 |
| Yes |  |  | 10.31 (6.01 - 17.67) | 10.08 (5.86 - 17.34) |
| SLAM service contact over<br>index period** |  |  |  |  |
| < 4 days |  |  | 1.00 | 1.00 |
| 4 to 7 days |  |  | 2.58 (1.02 - 6.53) | 2.62 (1.03 - 6.65) |
| 8 days and more |  |  | 3.56 (1.41 - 9.01) | 3.64 (1.42 - 9.30) |
| Smoking status |  |  |  |  |
| Current smoker |  |  |  | 1.00 |
| Past smoker |  |  |  | 1.06 (0.62 - 1.81) |
| never smoked |  |  |  | 0.69 (0.16 - 2.88) |
| BMI |  |  |  |  |
| Underweight & healthy |  |  |  | 1.00 |
| Overweight |  |  |  | 0.93 (0.49 - 1.75) |
| Obese |  |  |  | 1.86 (1.11 - 3.14) |

\*: first date of follow period is March 1, 2020
\*\*:: index period is between December 1, 2019 and March 1, 2020, which is 3 months prior to the follow-up period

## DISCUSSION

### Summary of findings

Our findings suggest that receiving clozapine treatment is associated with increased COVID-19 risk, compared to receiving any other type of antipsychotic treatment. Crude associations were attenuated but not completely explained by differences in sociodemographic factors such as age, gender, and ethnicity, factors related to health conditions such as smoking status, BMI, or proxies of availability of COVID testing (inpatient status or number of contacts with the SLAM services).

### Comparison with previous studies

To our knowledge, no previous research has specifically investigated the associations between infection with COVID-19 and receiving clozapine treatment, as compared to receiving treatment with other antipsychotics.

In previous research, the risk of COVID-19 has been reported to be associated with older age, male sex, ethnicity [having an African, Caribbean, Other Black background, Bangladeshi or Pakistani background, or Indian (if male)], and higher BMI (24,25). We found that older age was associated with COVID-19 infection and that infection rates were higher among Black people (compared to White people), and among people with high BMI (obese), but there were no significant associations with gender in our investigation.

### Strengths

The cohort was large and inclusive of all patients who met the inclusion criteria in a defined population. SLAM is a near-monopoly provider for all aspects of secondary mental health to a defined catchment area, so the study represents an almost comprehensive coverage of patients receiving clozapine treatment living in this catchment area of 1.3 million people.

In this analysis, the CRIS database made it possible to explore the complete electronic clinical records of more than six thousand patients who met our inclusion criteria, which gave us the statistical power to be able to analyse a relatively rare event, and adjust for a range of potential confounders.

In cohort studies, it is often impossible to be certain that the cases identified are true incident cases as opposed to prevalent cases that are identified during the study period. However, because there had been no cases of COVID in SLAM at the start of the follow-up period, we can be certain that these are all incident cases of COVID-19. Furthermore, we can completely rule out reverse causation: the prescription of clozapine could not have been affected by knowledge of COVID-19 status since clozapine status was measured before any cases of COVID had been diagnosed. Similarly, contact with services and inpatient status were measured before the start of the epidemic so could not have been affected by COVID-19 status.

### Limitations

We controlled for a number of potential confounders; however, there may still be residual confounding. There is a very large effect of inpatient status on the risk of COVID-19 infection. This is likely to arise partly from a higher risk of exposure to COVID-19 in hospital settings, and largely from the policy that inpatients showing any symptoms of COVID-19 were tested, while testing in the community was less comprehensive. Controlling for inpatient status on March 1, 2020, has not annulled the significant association between clozapine and COVID-19. However, we cannot rule out the possibility that clozapine-treated patients could be more likely to be tested for COVID-19, even after accounting for the differences in patient contact and inpatient status between the groups before March 1, 2020. During the study period, the Trust enacted a policy of attempting to discharge patients back into the community where possible, to free up inpatient capacity. We are making an assumption that the proportion of patients discharged did not differ between the clozapine treated group and the non-clozapine-treated group, and that the amount of care and monitoring before versus during the pandemic, remained proportional between groups.

The most recent BMI of some patients in the study were from almost 15 years ago. While this is likely to give some indication of the BMI of the patients, it is important to note that BMI is more likely to have been recently measured in clozapine-treated patients due to the increased monitoring.

The “current smoker” status of smoking data was extracted based on the status within a year prior to the follow-up period, so some that data was from almost a year ago. Some of the “past smoker” and “never smoked” data were from almost 18 years ago. Given the impact of smoking on clozapine metabolization and clozapine plasma levels, we cannot rule out that clozapine-treated patients may be questioned more frequently about smoking and therefore have more up-to-date information regarding smoking habits.

### Implications

To our knowledge, our results are the first to suggest that patients on clozapine treatment are at higher risk of infection by COVID-19 (21). This is consistent with previous research demonstrating that patients treated with clozapine have higher rates of infection and pneumonia than patients on other antipsychotics, and have alterations in both innate and adaptive immunity. There are also several alternative explanations for these findings, most notably the fact that clozapine-treated patients are likely to come into greater contact with services than patients on other antipsychotics and are therefore more likely to be tested if they develop symptoms. We have tried to adjust for patient contact, but, given the very large association between inpatient status and infection with COVID-19, we cannot confidently exclude the possibility that the association is explained by residual confounding.

The study is based on a relatively small number of cases, and we would not advocate any change in practice based on these findings alone. However, if the association is replicated and becomes firmly established, clinicians and patients will need to weigh up the increased risk of COVID-19 infection against the risk of psychotic relapse if clozapine is discontinued. Given that, for many patients, clozapine is the only effective antipsychotic, and with the well-established association between clozapine treatment and reduced all-cause mortality, these decisions are likely to be finely balanced and must be taken on a case by case basis.

Until this association is more firmly established, we would recommend that clinicians follow consensus guidelines for clozapine treatment during the COVID-19 pandemic, such as those of Siskind and colleagues (21,26). There should also be a focus on ensuring that clozapine-treated patients follow simple hygiene measures that can be taken to reduce the risks of COVID in clozapine-treated patients, including handwashing, social distancing and the rigorous use of facemasks and other personal protective equipment in clinical settings.

### Future research

As the COVID-19 pandemic progresses, we and other groups will be able to study this association in larger samples and with perhaps with better control of confounding. It will also be important to establish whether, among patients with COVID-19, clozapine-treated patients are at differential risk of adverse outcomes such as hospitalisation, pneumonia, treatment in intensive care or ventilation.

## Data Availability

Data are from clinical records and can not be shared externally

## Ethics statement

The research was conducted under ethical approval reference 18/SC/0372 from Oxfordshire Research Ethics Committee C.

## REFERENCES

1. Siskind D, McCartney L, Goldschlager R, Kisely S. Clozapine v. first- and second-generation antipsychotics in treatment-refractory schizophrenia: systematic review and meta-analysis. Br J Psychiatry. 2016 Nov;209(5):385–92.

2. Cho J, Hayes RD, Jewell A, Kadra G, Shetty H, MacCabe JH, et al. Clozapine and all-cause mortality in treatment-resistant schizophrenia: a historical cohort study. Acta Psychiatr Scand. 2018 Dec 16;139(3):acps.12989.

3. Kesserwani J, Kadra G, Downs J, Shetty H, MacCabe JH, Taylor D, et al. Risk of readmission in patients with schizophrenia and schizoaffective disorder newly prescribed clozapine. J Psychopharmacol. 2019 Apr 8;33(4):449–58.

4. Wimberley T, MacCabe JH, Laursen TM, Sørensen HJ, Astrup A, Horsdal HT, et al. Mortality and self-harm in association with clozapine in treatment-resistant schizophrenia. Am J Psychiatry. 2017 Oct 1;174(10):990–8.

5. Hayes RD, Downs J, Chang CK, Jackson RG, Shetty H, Broadbent M, et al. The effect of clozapine on premature mortality: an assessment of clinical monitoring and other potential confounders. Schizophr Bull. 2015;41(3):644–55.

6. Hayes JF, Marston L, Walters K, King MB, Osborn DPJ. Mortality gap for people with bipolar disorder and schizophrenia: UK-based cohort study 2000-2014. Vol. 211, British Journal of Psychiatry. Royal College of Psychiatrists; 2017. p. 175–81.

7. Vermeulen JM, Van Rooijen G, Van De Kerkhof MPJ, Sutterland AL, Correll CU, De Haan L. Clozapine and Long-Term Mortality Risk in Patients with Schizophrenia: A Systematic Review and Meta-analysis of Studies Lasting 1.1-12.5 Years. Vol. 45, Schizophrenia Bulletin. Oxford University Press; 2019. p. 315–29.

8. Chou FHC, Tsai KY, Chou YM. The incidence and all-cause mortality of pneumonia in patients with schizophrenia: A nine-year follow-up study. J Psychiatr Res. 2013;47(4):460–6.

9. Seminog OO, Goldacre MJ. Risk of pneumonia and pneumococcal disease in people with severe mental illness: English record linkage studies. Thorax. 2013;68(2):171–6.

10. Shen TC, Chen CH, Huang YJ, Lin CL, Chang TC, Tu CY, et al. Risk of pleural empyema in patients with schizophrenia: A nationwide propensity-matched cohort study in Taiwan. BMJ Open. 2018 Jul 1;8(7).

11. John A, McGregor J, Jones I, Lee SC, Walters JTR, Owen MJ, et al. Premature mortality among people with severe mental illness - New evidence from linked primary care data. Schizophr Res. 2018 Sep 1;199:154–62.

12. Gurillo P, Jauhar S, Murray RM, MacCabe JH. Does tobacco use cause psychosis? Systematic review and meta-analysis. The Lancet Psychiatry. 2015 Aug 1;2(8):718–25.

13. Haddad PM. Current use of second-generation antipsychotics may increase risk of pneumonia in people with schizophrenia. Evid Based Ment Health. 2013 Nov;16(4):109.

14. Kuo C-J, Yang S-Y, Liao Y-T, Chen WJ, Lee W-C, Shau W-Y, et al. Second-generation antipsychotic medications and risk of pneumonia in schizophrenia. Schizophr Bull. 2013 May;39(3):648–57.

15. Stoecker ZR, George WT, O’Brien JB, Jancik J, Colon E, Rasimas JJ. Clozapine usage increases the incidence of pneumonia compared with risperidone and the general population: A retrospective comparison of clozapine, risperidone, and the general population in a single hospital over 25 months. Int Clin Psychopharmacol. 2017;32(3):155–60.

16. De Leon J, Sanz EJ, De las Cuevas C. Data From the World Health Organization’s Pharmacovigilance Database Supports the Prominent Role of Pneumonia in Mortality Associated With Clozapine Adverse Drug Reactions. Schizophr Bull. 2020;46(1):1–3.

17. Liu NH, Daumit GL, Dua T, Aquila R, Charlson F, Cuijpers P, et al. Excess mortality in persons with severe mental disorders: a multilevel intervention framework and priorities for clinical practice, policy and research agendas. World Psychiatry. 2017 Feb 1;16(1):30–40.

18. Gurrera RJ, Perry NL. Clozapine-Associated Aspiration Pneumonia: Case Series and Review of the Literature: Reply. Vol. 60, Psychosomatics. Elsevier Inc.; 2019. p. 103.

19. Li XH, Zhong XM, Lu L, Zheng W, Wang S Bin, Rao WW, et al. The prevalence of agranulocytosis and related death in clozapine-treated patients: A comprehensive meta-analysis of observational studies. Psychol Med. 2020 Mar 1;50(4):583–94.

20. Ponsford M, Castle D, Tahir T, Robinson R, Wade W, Steven R, et al. Clozapine is associated with secondary antibody deficiency. Br J Psychiatry. 2019 Jan 1;214(2):83–9.

21. Siskind D, Honer WG, Clark S, Correll CU, Hasan A, Howes O, et al. Consensus statement on the use of clozapine during the COVID-19 pandemic. J Psychiatry Neurosci. 2020 Apr 3;45(3):2.

22. Stewart R, Soremekun M, Perera G, Broadbent M, Callard F, Denis M, et al. The South London and Maudsley NHS Foundation Trust Biomedical Research Centre (SLAM BRC) case register: Development and descriptive data. BMC Psychiatry. 2009 Aug 12;9(1):51.

23. CRIS NLP Applications Library. CRIS Natural Language Processing [Internet]. v1.1. 2020. Available from: https://www.maudsleybrc.nihr.ac.uk/facilities/clinical-record-interactive-search-cris/cris-natural-language-processing/

24. Liu K, Chen Y, Lin R, Han K. Clinical features of COVID-19 in elderly patients: A comparison with young and middle-aged patients. Journal of Infection. W.B. Saunders Ltd; 2020.

25. White C, Nafilyan V. Coronavirus (COVID-19) related deaths by ethnic group, England and Wales - Office for National Statistics [Internet]. 2020. Available from: https://www.ons.gov.uk/peoplepopulationandcommunity/birthsdeathsandmarriages/deaths/articles/coronavirusrelateddeathsbyethnicgroupenglandandwales/2march2020to10april2020

26. Luykx JJ, van Veen SMP, Risselada A, Naarding P, Tijdink JK, Vinkers C. Safe and informed prescribing of psychotropic medication during the COVID-19 pandemic. Br J Psychiatry. 2020 May 4;1–9.

